# Exploring the impact of an artificial intelligence-based intraoperative image navigation system in laparoscopic surgery on clinical outcomes: A protocol for a multicenter randomized controlled trial

**DOI:** 10.1101/2024.08.05.24310603

**Authors:** Daichi Kitaguchi, Nozomu Fuse, Masashi Wakabayashi, Norihito Kosugi, Yuto Ishikawa, Kazuyuki Hayashi, Hiro Hasegawa, Nobuyoshi Takeshita, Masaaki Ito

## Abstract

**Background:** In the research field of artificial intelligence (AI) in surgery, there are many open questions that must be clarified. Well-designed randomized controlled trials (RCTs) are required to explore the positive clinical impacts by comparing the use and non-use of AI-based intraoperative image navigation. Therefore, herein, we propose the “ImNavi” trial, a multicenter RCT, to compare the use and non-use of an AI-based intraoperative image navigation system in laparoscopic surgery.

**Methods:** The ImNavi trial is a Japanese multicenter RCT involving 1:1 randomization between the use and non-use of an AI-based intraoperative image navigation system in laparoscopic colorectal surgery. The participating institutions will include three high-volume centers with sufficient laparoscopic colorectal surgery caseloads (>100 cases/year), including one national cancer center and two university hospitals in Japan. Written informed consent will be obtained from all patients. Patients aged between 18 and 80 years scheduled to undergo laparoscopic left-sided colorectal resection will be included in the study. The primary outcome is the time required for each target organ, including the ureter and autonomic nerves, to be recognized by the surgeon after its initial appearance on the monitor. Secondary outcomes include intraoperative target organ injuries, intraoperative complications, operation time, blood loss, duration of postoperative hospital stay, postoperative complications within 30 days, postoperative male sexual dysfunction 1 month after surgery, surgeon’s confidence in recognizing each target organ, and the postoperative fatigue of the primary surgeon.

**Discussion:** The impact of AI-based surgical applications on clinical outcomes beyond numerical expression will be explored from a variety of viewpoints while evaluating quantitative items, including intraoperative complications and operation time, as secondary endpoints. We expect that the findings of this RCT will contribute to advancing research in the domain of AI in surgery.

**Trial registration:** The trial was registered at the University Hospital Medical Information Network Center (https://www.umin.ac.jp/ctr/index-j.html) on March 28th, 2023 under trial ID: UMIN000050701.

## Background

Surgical volumes are large and show an increasing trend worldwide [1]; nevertheless, it is estimated that 143 million additional surgical procedures are required annually to save lives and prevent disabilities [2]. Surgery is an essential aspect of healthcare and is associated with increased life expectancy [1]; however, inter-institutional disparities in surgical skill levels remain. The introduction of minimally invasive surgery (MIS), including endoscopic surgical approaches, has made surgery more effective [3]. However, surgery has become more complex, and achieving basic surgical skill levels, which are directly linked to postoperative clinical outcomes, has become more difficult [4, 5]. Improvements in the safety and efficiency of surgery must be a major component in strengthening healthcare systems, and technological innovation may be one of the solutions to achieve this goal.

Computer vision (CV) based on artificial intelligence (AI), particularly deep learning using convolutional neural networks, is a recent technological innovation. AI-based CV is a field of computer science that enables AI to extract meaningful information from digital images and videos, and to process and make recommendations based on that information [6]. In recent years, AI-based CV has achieved remarkable success in image recognition tasks in the field of medical image diagnosis, such as radiology [7–9], pathology [10, 11], gastroenterology [12], and ophthalmology [13–15]. In the field of surgery, several AI-based CV applications for MIS, specifically semantic segmentation-based intraoperative image navigation, have been developed based on the nature of MIS, which relies heavily on the visual information provided by surgical endoscopes [16–18]. These are expected to reduce intraoperative anatomical cognitive errors. However, the majority of studies in this field are still in the proof-of-concept stage.

In the research field of AI in surgery, many open questions regarding how AI-based intraoperative image navigation affects surgeon performance exist, including whether it can achieve some improvement in clinical outcomes, and whether it works properly in diverse environments without overfitting. To address these questions, well-designed randomized controlled trials (RCTs) are required to explore the positive clinical impacts of AI-based intraoperative image navigation. Therefore, we propose the “ImNavi” trial, a multicenter RCT, to compare the use and non-use of an AI-based intraoperative image navigation system in laparoscopic surgery.

## Methods/Design

### Participants, interventions, and outcomes

#### Trial design

The ImNavi trial is designed as a Japanese multicenter RCT with 1:1 randomization between the use and non-use of an AI-based intraoperative image-navigation system in laparoscopic surgery. The study protocol was prepared according to the reporting guidelines of the Standard Protocol Items: Recommendations for Interventional Trials - Artificial Intelligence extension (SPIRIT-AI) [19]. The SPIRIT-AI checklist is presented in Additional File 1.

#### Study setting and recruitment

This hospital-based study will be conducted in Japan. Eligible candidates will be identified from patients referred to a colorectal surgeon or colorectal cancer multidisciplinary team based on colonoscopy, computed tomography, or magnetic resonance imaging findings, and eligibility will be confirmed after a review of the study criteria. The target surgical procedures in this study are left-sided colorectal resections, including left hemicolectomy, sigmoidectomy, anterior resection, intersphincteric resection, Hartmann’s procedure, and abdominoperineal resection. The target organs for semantic segmentation are the ureter and autonomic nerves, including the hypogastric nerves and the aortic plexus. The AI-based intraoperative image navigation system used in this study is based on an existing semantic segmentation algorithm that we developed [20, 21]. The accuracy and performance of both the ureter recognition model (UreterNet) and the autonomic nerve recognition model (NerveNet) have been validated in our previous study [20]. Alienware x15 R2 (Dell Technologies Inc., Round Rock, Texas, U.S.) was used as the computational system in this study.

#### Eligibility criteria

Patients aged between 18 and 80 years scheduled to undergo laparoscopic left-sided colorectal resection and who provided written informed consent will be included in the ImNavi trial. Long-term outcomes will not be included in the endpoints; therefore, patients with any colorectal disease will be eligible. Patients with emergent surgery, expected severe intra-abdominal adhesion, significant anatomical anomaly, and those deemed unsuitable by the doctor in charge will be excluded. The trial flow diagram is illustrated in Figure 1.

**Figure 1:**
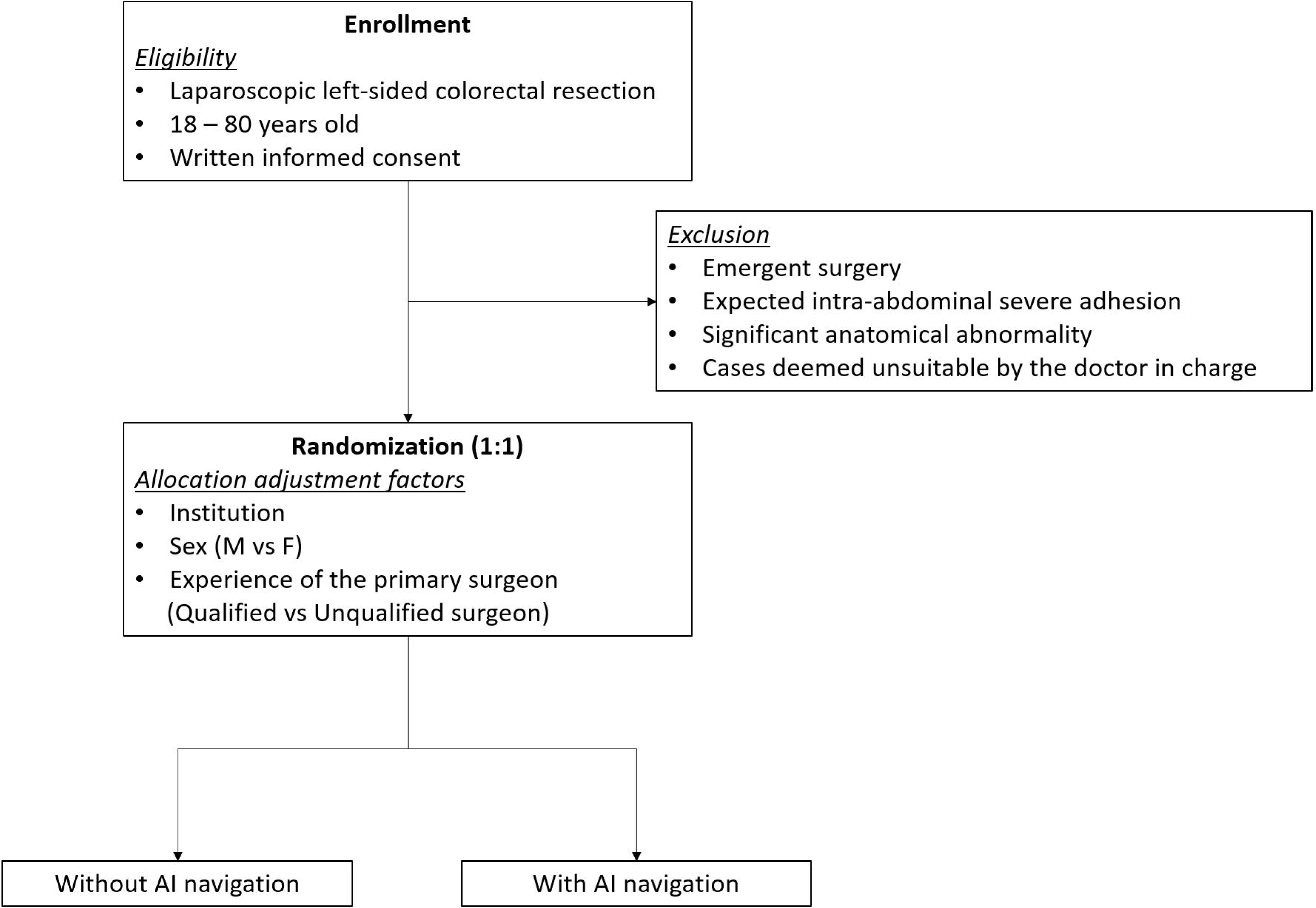
Trial flow diagram.

#### Interventions

The time required to recognize each target organ, including the ureter and autonomic nerves, after its initial appearance on the monitor will be compared between the surgeons with and without an AI-based intraoperative image navigation system. The initial time of appearance of each target organ will be retrospectively determined by two or more judges observing via video. The system output, semantic segmentation pixels, will be superimposed on a sub-monitor placed adjacent to the main monitor, as illustrated in Figure 2, without influencing the display of the main monitor. The surgeons will decide the time and frequency of viewing the sub-monitor. The primary surgeon will be asked to orally state that they recognize each target organ during surgery. All intraoperative statements made by the primary surgeon will be recorded by a microphone placed near the operating table. The intraoperative audio will be collated with the intraoperative video, and the difference in time points between the initial appearance on the monitor and the primary surgeon’s recognition (vocalization) will be calculated for each target organ.

**Figure 2:**
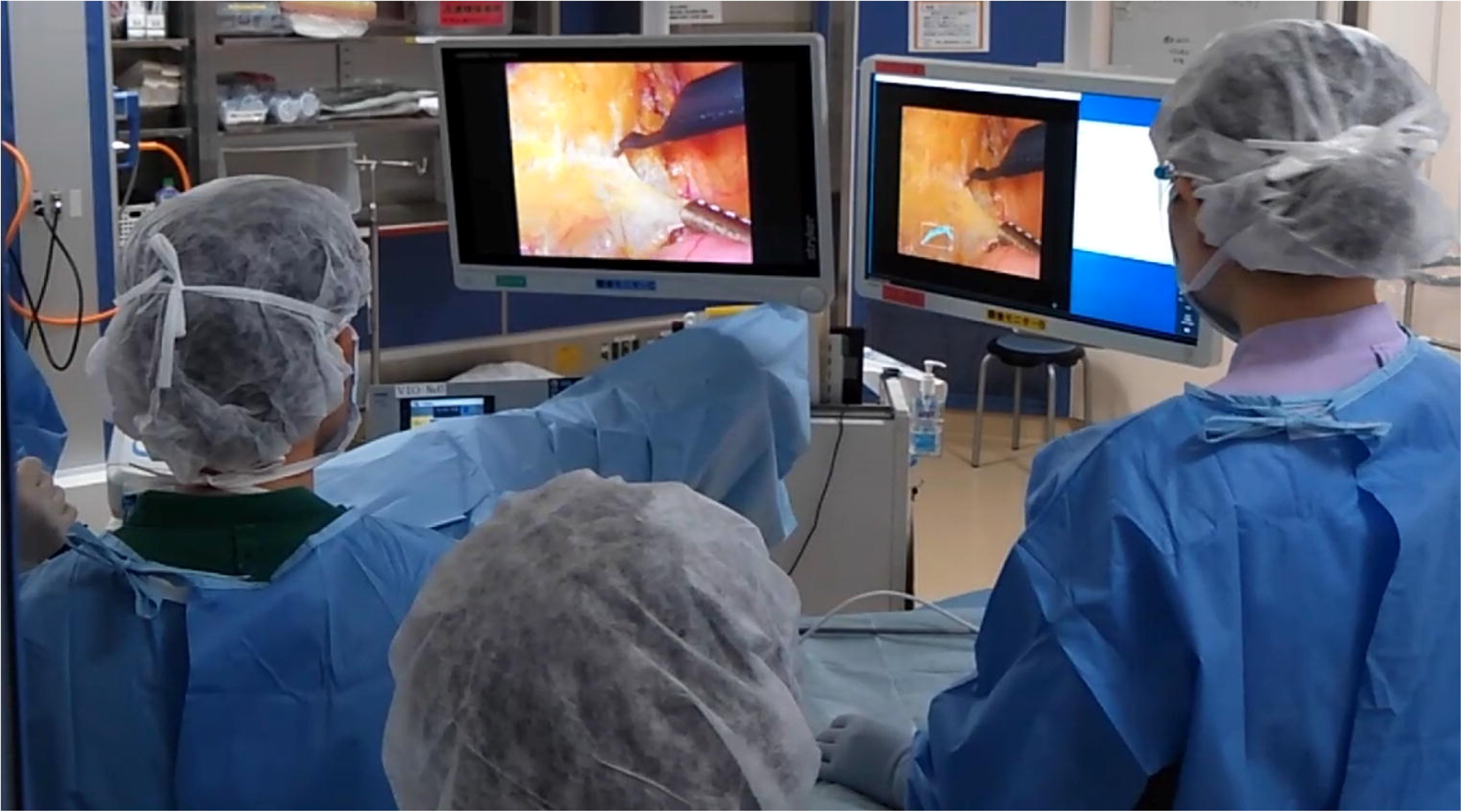
Example of the monitor layout in an operation room. The system output (semantic segmentation pixels) is superimposed on a sub-monitor placed adjacent to the main monitor, and does not affect the main monitor.

#### Outcomes

The primary outcome is the time required for each target organ, including the ureter and autonomic nerves, to be recognized by the surgeon after its initial appearance on the monitor. Secondary outcomes include intraoperative target organ injuries; intraoperative complications; total operation time; operation time for each surgical step divided by existing definitions [22]; blood loss; duration of postoperative hospital stay; postoperative complications within 30 days; postoperative male sexual dysfunction 1 month post-surgery; the surgeon’s confidence in recognising each target organ; and postoperative fatigue of the primary surgeon, according to the Piper Fatigue Scale-12 (PFS-12) [23]. The severity of all intraoperative and postoperative complications, according to the Common Terminology Criteria for Adverse Events and the Clavien–Dindo classification, will also be collected as secondary outcomes.

#### Participant timelines

Before surgery, written informed consent will be obtained from all patients and their primary surgeons, and patients will be allocated to their respective groups using an electronic data capture (EDC) system. A preoperative report, including patient, disease, and primary surgeon background information, will be provided using case report forms (CRFs). After surgery, an intraoperative report on the primary surgeon’s surgical information and postoperative fatigue, according to the PFS-12, will be provided using CRFs and questionnaires, respectively. One month after surgery, the postoperative course and outcome reports, including postoperative complications and date of discharge, will be provided using the CRFs. This study will not regulate modalities or intervals between examinations for each participating institution, or intervene in each institution’s routine clinical practice.

#### Sample size

Based on non-published data from previous studies at our institution, the mean time (± standard deviation) required by surgeons to recognize each target organ without using the AI-based intraoperative image navigation system after its initial appearance on the monitor is estimated to be 6.0 (± 9.0) s. To detect a 2-s decrease with the AI-based system, using a power of 80% and a one-sided significance level of 0.15 in the Wilcoxon rank-sum test, 38 patients in each group (76 patients in total) would be required for this RCT. After accounting for dropouts and ineligibility, an additional 14 patients will be included. Thus, the target sample size would be 90 patients.

#### Assignment of interventions

Once eligibility is established, patients will be allocated to either the use or non-use of AI-based intraoperative image navigation groups. Randomization, performed by computers through the Internet (https://www2.epoc-ncc.net/), will be adjusted using the minimization method with a random component to balance the groups, participating institution, sex (male versus female), and experience of the primary surgeon (qualified versus unqualified by the Endoscopic Surgical Skill Qualification System of the Japan Society for Endoscopic Surgery) [24]. Patients will be randomized in a 1:1 ratio. Data will be analyzed on an ‘intention-to-treat’ basis when patients are not subjected to a randomized treatment modality.

### Data collection, management, and analysis

Data collection will be carried out using CRFs and a validated PFS-12 questionnaire. All data will be entered into an EDC system. Regular data quality checks will be performed annually in accordance with the Quality Management Plan. All data will be handled following the Ethical Guidelines for Medical and Biological Research Involving Human Subjects. Data backups will be stored in secure fireproof locations, and test restorations will be performed regularly. After completion of the trial, all essential trial documentation and source documents, including signed informed consent forms and copies of CRFs, will be securely retained for at least 5 years.

#### Statistical analysis

Baseline numerical data will be described as the mean (± standard deviation) or median (range). Baseline categorical data will be presented as percentages. All comparative analyses will be conducted on an ‘intention-to-treat’ basis. In addition to the intention-to-treat analysis, per-protocol and as-treated analyses will be applied as sensitivity analyses. Primary outcomes will be compared using the Wilcoxon’s rank sum test. Secondary outcomes will be compared using the Fisher’s exact test or the Wilcoxon’s rank sum test, as necessary.

#### Monitoring

Regular data quality checks will be performed yearly, per the Quality Management Plan. Central monitoring will be conducted by the data center based on the CRF data collected via the EDC system. The monitoring staff will prepare a monitoring report after central monitoring and report it to the principal investigator.

### Ethics and dissemination

The participating institutions will include three high-volume centers with sufficient laparoscopic colorectal surgery caseloads (>100 cases/year), including one national cancer center and two university hospitals in Japan. The protocol for this RCT has been reviewed and approved by the ethics committees of each participating institution, and the trial has been registered at University Hospital Medical Information Network Center (https://www.umin.ac.jp/ctr/index-j.html) under the trial ID UMIN000050701. Written informed consent will be obtained from all patients after a thorough oral explanation by the doctor in charge at each center participating in the ImNavi trial. All procedures will be conducted in accordance with the ethical standards of the Declaration of Helsinki.

## Discussion

Recently, analogous digital solutions have been translated and clinically implemented for diagnostic applications in gastrointestinal endoscopy [25] and radiology [26]. In the field of surgery, several deep learning-based CV solutions have been developed by academic and industry groups, mostly for MIS. Significant work has also been performed on potential use cases [3]; however, no CV tools are widely used for diagnostic or therapeutic applications in surgery. Recently, an intraoperative AI system identifying anatomical landmarks for laparoscopic cholecystectomy has been developed and reported [18]; similarly, in the urological field, an AI alert system that can predict the occurrence of intraoperative bleeding events during robotic surgery has been developed [27]. Although both systems could potentially be used as tools to improve the safety of the intervention, they were only tested in a trial involving 10 patients at a single center. Establishing solid evidence through RCTs is essential before AI can be applied in surgery to move beyond the proof-of-concept step to clinical implementation. In addition, a multicenter study design is desirable to eliminate bias due to overfitting and to demonstrate generalization performance. This study is the first multicenter RCT to be designed to explore the clinical significance of AI-based intraoperative image navigation.

The target CV task of this study is the semantic segmentation of the ureter and autonomic nerves in laparoscopic colorectal surgery. Iatrogenic ureteral injury in colorectal surgery is a rare but potentially devastating complication of colorectal surgery. The incidence is approximately 0.5%–1.5% [28–30]; however, a significant increase has been observed with laparoscopy compared to open colectomies [29, 30] due to the lack of tactile information. In addition, trauma to the aortic plexus may occur during high ligation of the inferior mesenteric artery, and the superior hypogastric nerves may be injured along the sacral promontory or presacral region [31]. Injuries to the sympathetic nerves at this location affect the ability to ejaculate, including retrograde ejaculation, whereas injuries to the parasympathetic nerves result in erectile dysfunction [31]. A previous study reported a decrease in male sexual function following colorectal surgery, with a decrease from 78 to 32% in ejaculatory function, 71 to 24% in erectile function, and 82 to 57% in sexual activity [32]; these dysfunctions are often overlooked but should be strictly avoided.

The time required for recognition of each target organ by surgeons after its initial appearance on the monitor was set as the primary endpoint of this study, with the expectation that the AI-based intraoperative image navigation system would contribute to the more rapid recognition of target organs. Intraoperative target organ injury is a more straightforward primary endpoint; however, the number of events was considered too small for analysis. In addition, we believe that more rapid recognition of key anatomical landmarks during surgery can lead to several benefits other than prevention of organ injury, such as identification of the correct dissection plane, and confidence in proceeding with the procedure. The impact of AI-based surgical applications on clinical outcomes beyond numerical expression will be explored from various facets while evaluating quantitative items, including intraoperative complications and operation time, as secondary endpoints. Furthermore, we expect that the findings of this RCT will contribute to advancing research in the domain of AI in surgery.

## Data Availability

All data produced in the present work are contained in the manuscript

## Strengths and limitations of this study

- The ImNavi trial is a Japanese multicenter RCT with 1:1 randomization between the use and non-use of an artificial intelligence-based intraoperative image-navigation system in laparoscopic surgery.
- Eligible candidates will be identified from patients referred to a colorectal surgeon or colorectal cancer multidisciplinary team based on colonoscopy, computed tomography, or magnetic resonance imaging findings, and eligibility will be confirmed following a review of the criteria.
- The primary outcome is the time required for each target organ to be recognized by the surgeon after its initial appearance on the monitor.
- Secondary outcomes include intraoperative target organ injuries; intraoperative complications; total operation time; operation time for each surgical step divided by existing definitions; blood loss; duration of postoperative hospital stay; postoperative complications within 30 days; postoperative male sexual dysfunction 1 month after surgery; surgeon’s confidence in recognition of each target organ; and postoperative fatigue of the primary surgeon, according to the Piper Fatigue Scale-12.
- The severity of all intraoperative and postoperative complications, measured according to Common Terminology Criteria for Adverse Events and the Clavien–Dindo classification, will also be collected as secondary outcomes.

## Trial Status

This trial represents the first version of the study protocol. This trial was registered at the University Hospital Medical Information Network Center (https://www.umin.ac.jp/ctr/index-j.html) on March 28th, 2023 (trial ID: UMIN000050701). Recruitment will begin on June 2023 and is scheduled to be completed by December 2024.

## LIST OF ABBREVIATIONS

AI: artificial intelligence
RCT: randomized controlled trials
MIS: minimally invasive surgery
CV: computer vision
EDC: electronic data capture
CRF: case report forms

## DECLARATIONS

### Ethics approval and consent to participate

The protocol for this RCT has been reviewed and approved by the ethics committees of each participating institution, and the trial has been registered at the University Hospital Medical Information Network Center (https://www.umin.ac.jp/ctr/index-j.html) under the trial ID UMIN000050701. Written informed consent will be obtained from all patients after a thorough oral explanation by the doctor in charge at each center participating in the ImNavi trial. All procedures will be conducted in accordance with the ethical standards of the Declaration of Helsinki.

### Consent for publication

Not applicable

### Availability of data and materials

Not applicable

### Authors’ contributions

All authors made substantial contributions to the study concept or data analysis and interpretation; drafted the manuscript or revised it critically for important intellectual content; approved the final version of the manuscript to be published; and agreed to be accountable for all aspects of the work.

## Acknowledgements

We would like to thank all the members of the Clinical Research Support Office of the National Cancer Center Hospital East for their continuous support during the planning of this study.

## Additional files

**Additional file 1:** Ethical approval document (English translation version)

**Additional file 2:** Copy of the original funding documentation (English translation version)

**Additional file 3:** A completed SPIRIT checklist

**Additional file 4:** A SPIRIT figure

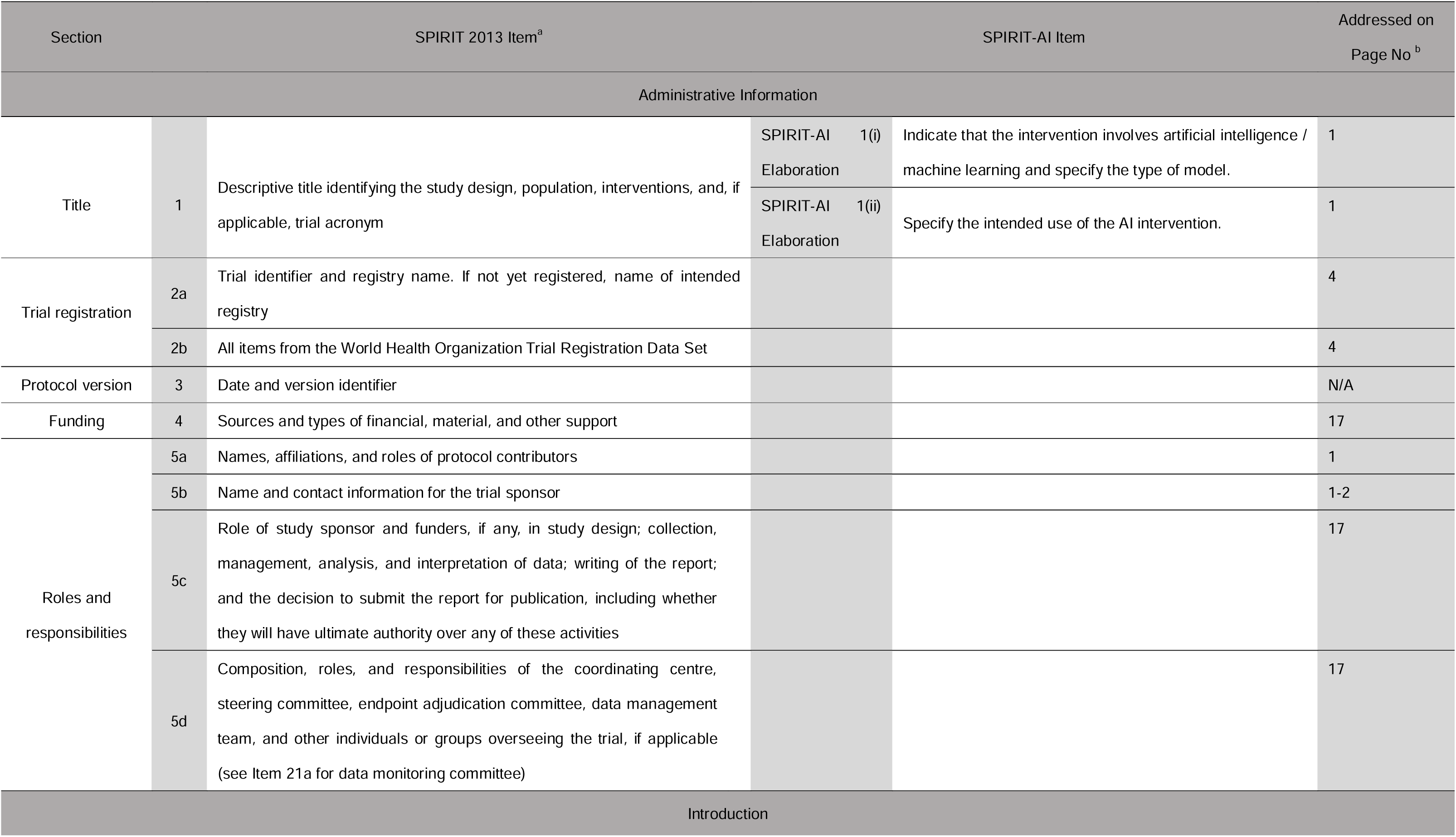
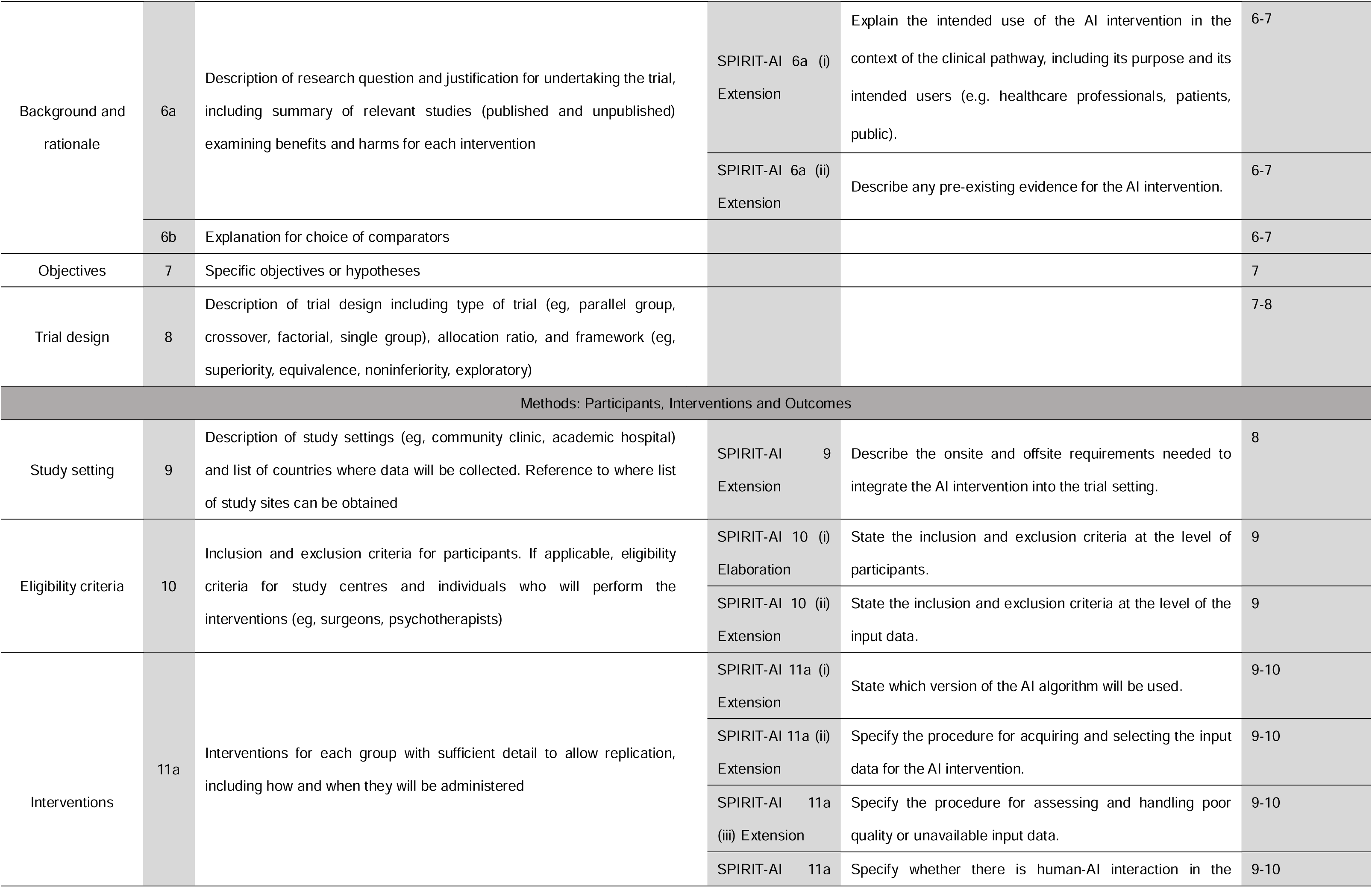
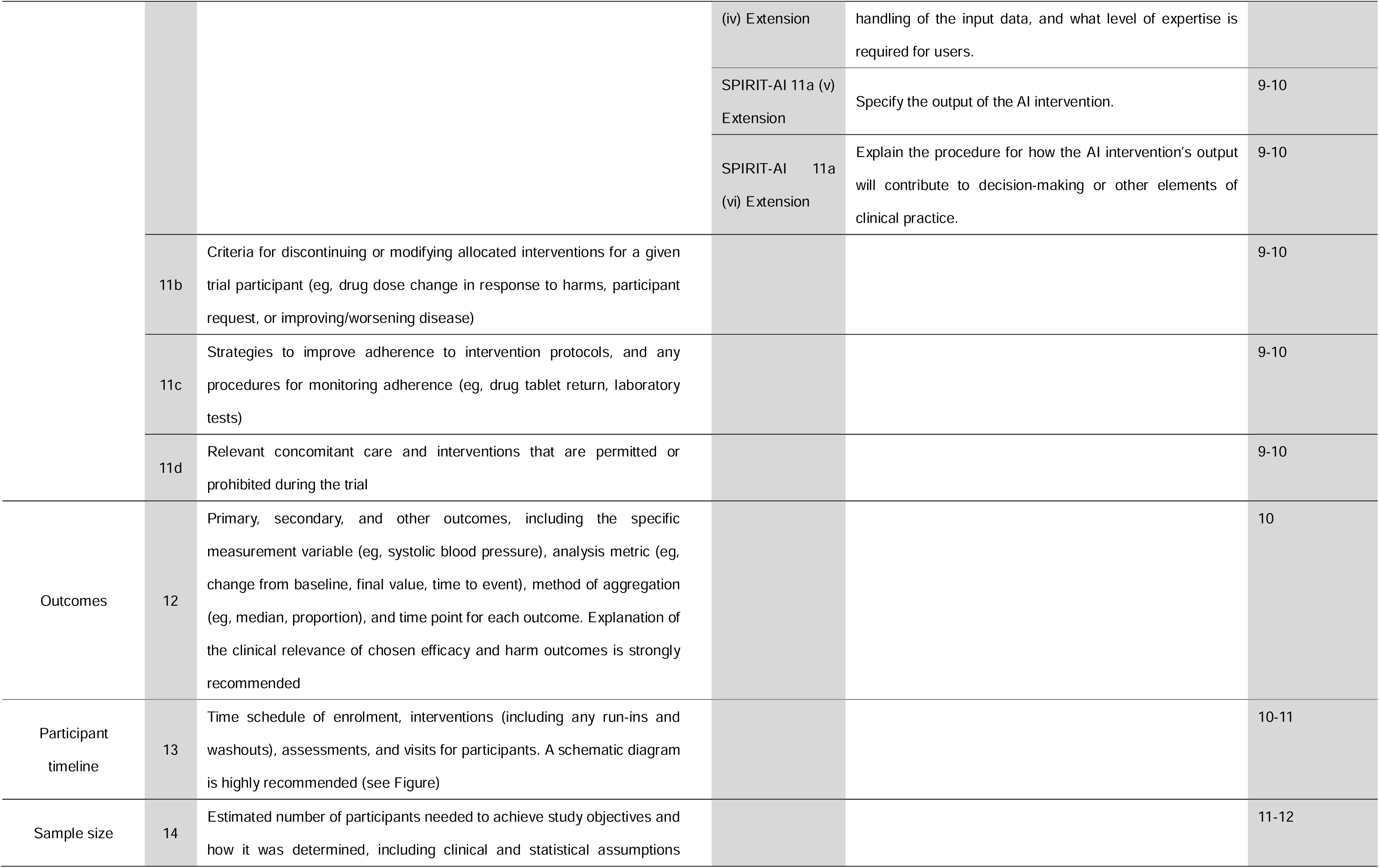
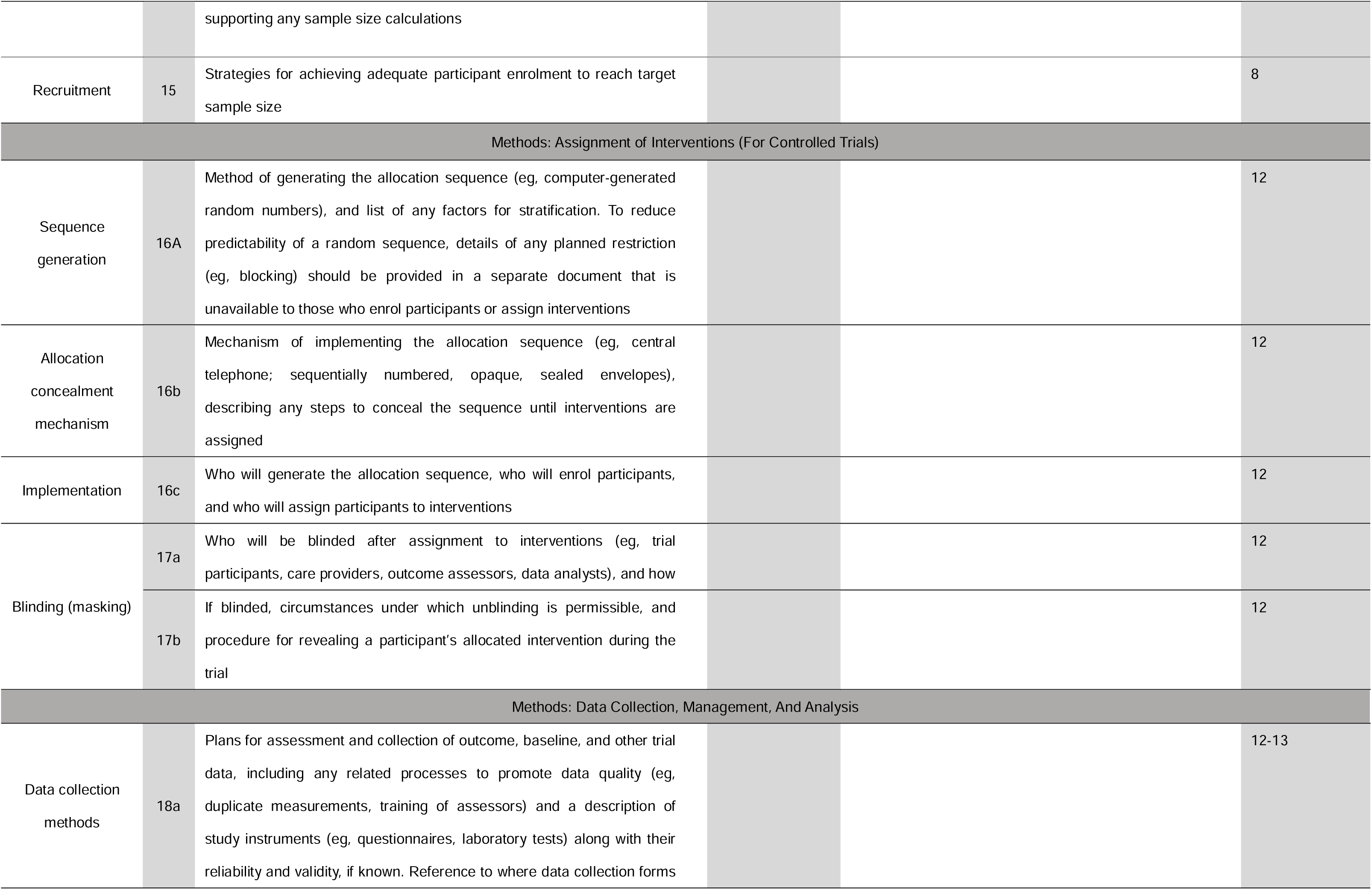
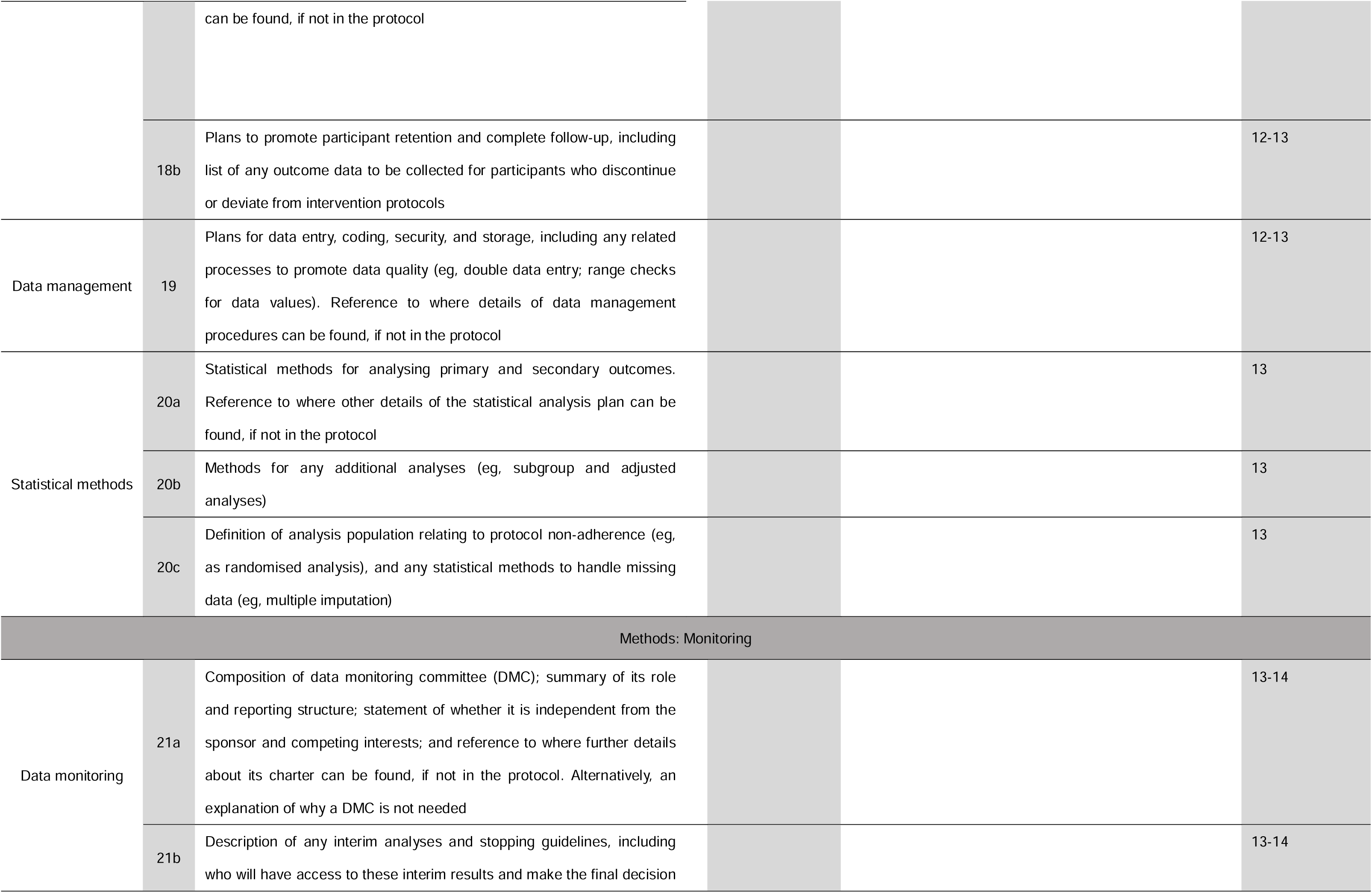
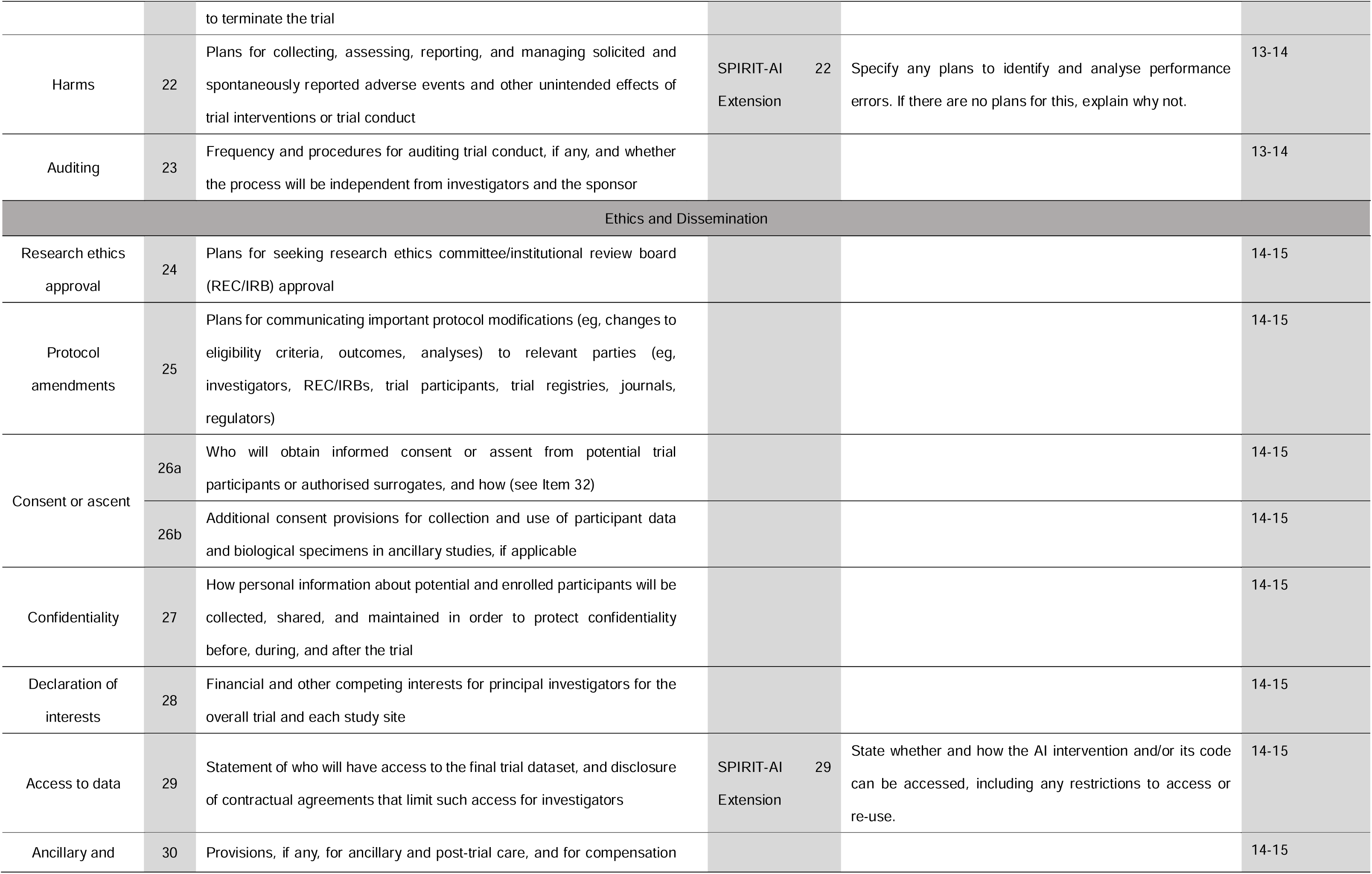
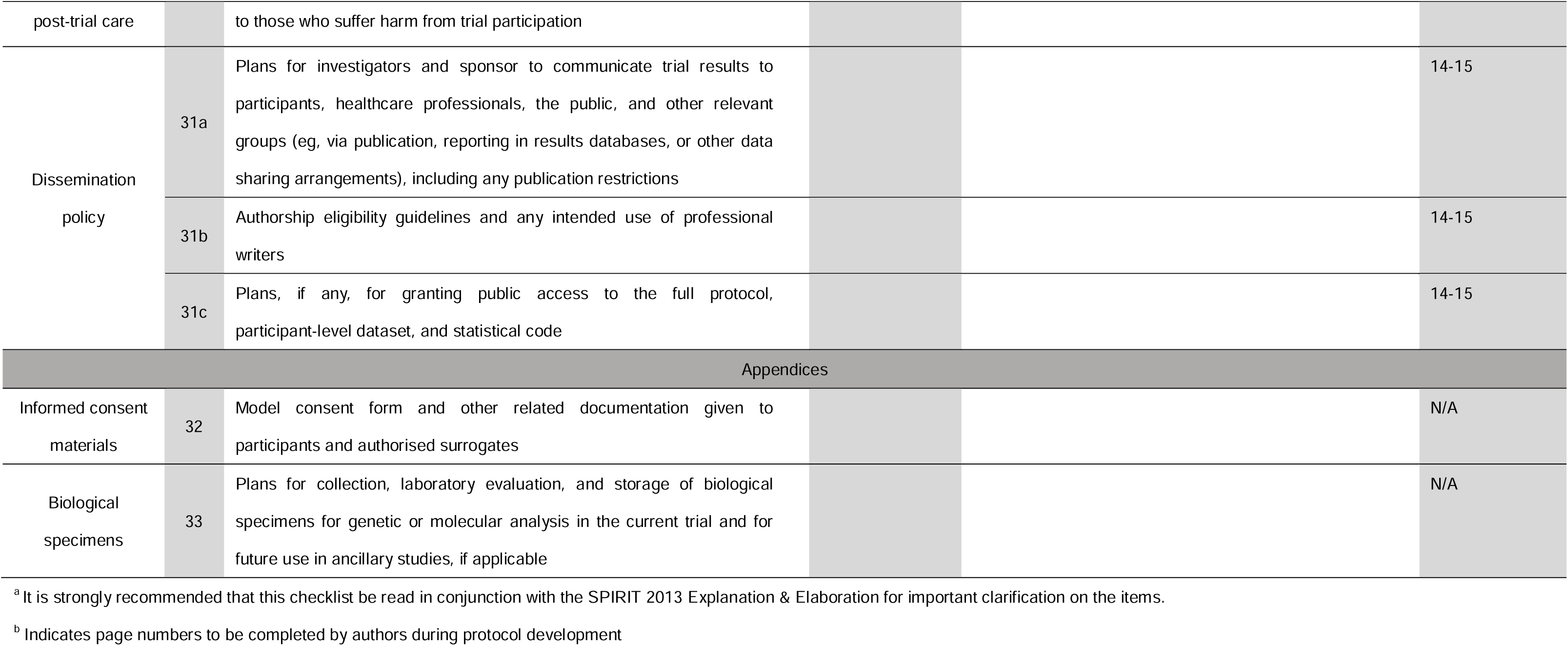
SPIRIT-AI Checklist: Recommended items to address in a protocol and related documents for clinical trials evaluating AI interventions.

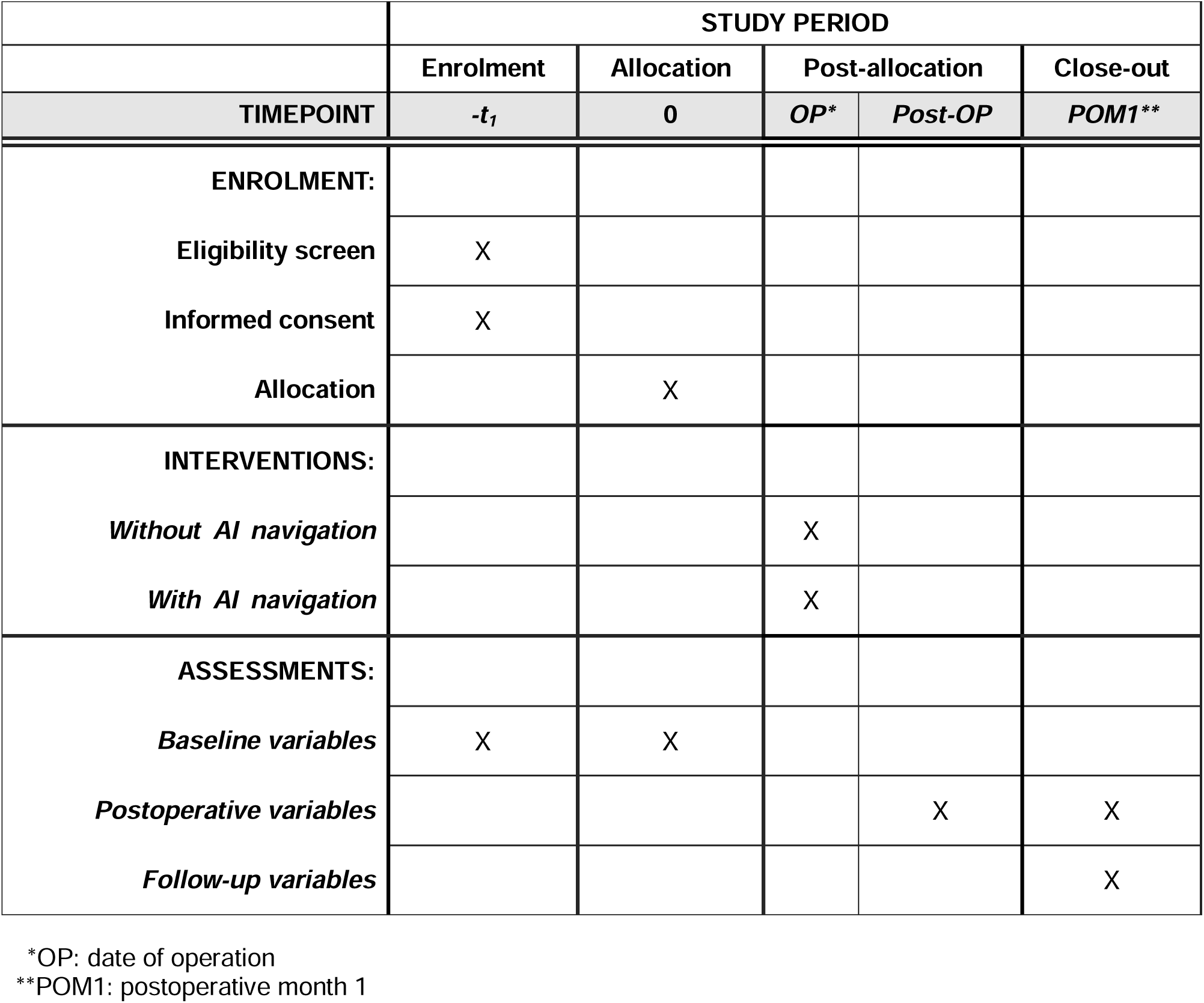
SPIRIT figure: The schedule of enrolment, interventions, and assessments.

